# Clinical Prediction Models for Primary Prevention of Cardiovascular Disease: Validity in Independent Cohorts

**DOI:** 10.1101/2021.01.31.21250871

**Authors:** Gaurav Gulati, Riley J Brazil, Jason Nelson, David van Klaveren, Christine M. Lundquist, Jinny G. Park, Hannah McGinnes, Ewout W. Steyerberg, Ben Van Calster, Benjamin S. Wessler, David M. Kent

## Abstract

**Background:** Clinical prediction models (CPMs) are used to inform treatment decisions for the primary prevention of cardiovascular disease. We aimed to assess the performance of such CPMs in fully independent cohorts.

**Methods and Results:** 63 models predicting outcomes for patients at risk of cardiovascular disease from the Tufts PACE CPM Registry were selected for external validation on publicly available data from up to 4 broadly inclusive primary prevention clinical trials. For each CPM-trial pair, we assessed model discrimination, calibration, and net benefit. Results were stratified based on the relatedness of derivation and validation cohorts, and net benefit was reassessed after updating model intercept, slope, or complete re-estimation. The median c statistic of the CPMs decreased from 0.77 (IQR 0.72-0.78) in the derivation cohorts to 0.63 (IQR 0.58-0.66) when externally validated. The validation c-statistic was higher when derivation and validation cohorts were considered related than when they were distantly related (0.67 vs 0.60, p < 0.001). The calibration slope was also higher in related cohorts than distantly related cohorts (0.69 vs 0.58, p < 0.001). Net benefit analysis suggested substantial likelihood of harm when models were externally applied, but this likelihood decreased after model updating.

**Conclusions:** Discrimination and calibration decrease significantly when CPMs for primary prevention of cardiovascular disease are tested in external populations, particularly when the population is only distantly related to the derivation population. Poorly calibrated predictions lead to poor decision making. Model updating can reduce the likelihood of harmful decision making, and is needed to realize the full potential of risk-based decision making in new settings.

## Introduction

Cardiovascular disease (CVD) is the leading cause of mortality in the United States. Interventions to reduce the incidence of cardiovascular disease in the primary prevention setting abound, including lifestyle interventions, antihypertensive therapy, lipid lowering therapy, and antiplatelet therapy.^1–4^ A key aspect of optimizing primary prevention strategies involves targeting interventions to high risk individuals, and some have argued that clinical trial results should be routinely interpreted using risk-based analyses.^5,6^

Clinical prediction models (CPMs) are critical in estimating individual patient risk and are being increasingly used to guide treatment decisions for primary prevention.^7^ In order for CPMs to be beneficial however, their performance must be robust to validation on new cohorts (external validation). It is known that in general, the performance of CPMs degrade when models are externally validated, both in terms of discrimination (distinguishing high and low risk individuals) and calibration (accuracy of risk estimates).^8^ This decrement in performance can significantly threaten the utility of many CPMs by leading to harmful treatment decisions if used naïvely in routine clinical practice. An illustrative example is that of the recent controversy surrounding the use of the Pooled Cohort Equations (PCE) in guiding primary prevention statin therapy. Guidelines recommend statin therapy for individuals with a PCE-estimated 10-year risk of CVD ≥ 7.5%,^9^ but once it was shown that the PCE systematically overestimated risk in several populations, potentially resulting in significant overtreatment, some argued that outcomes might be better using “trial-based” guidelines—which emphasize applying the best treatment on average to populations defined by trial inclusion and exclusion criteria—instead of “risk-based” guidelines.^10^

Despite the increasing number of CPMs predicting incident CVD in the literature, most have not been validated in external cohorts, and those that have are usually validated on a single external cohort.^11,12^ Therefore, the performance of CPMs for incident CVD when externally validated is largely unknown. Thus, the primary aim of this study was to characterize the performance of CPMs for incident CVD by performing independent external validations on patient-level data from publicly available clinical trial databases. We also assessed the impact of various model updating procedures on model performance.

## Methods

### Source of models

The Tufts Predictive Analytics and Comparative Effectiveness (PACE) CPM Registry is a registry of CPMs published between January 1990 and December 2015 that predict outcomes in patients at risk for or with known cardiovascular disease. Detailed methods for development of the registry have been reported previously.^11^ Briefly, for inclusion in the registry, articles must (1) develop a CPM as a primary aim, (2) contain at least 2 outcome predictors, and (3) present enough information to estimate the probability for an individual patient. For this analysis, we selected from the registry all CPMs predicting outcomes on healthy patients at risk for CVD.

### Source of validation cohorts

De-identified patient-level data from four clinical trials were obtained from the National Heart, Lung, and Blood Institute (NHLBI) via application to the Biologic Specimen and Data Repository Information Coordinating Center (BioLINCC). The trials used in this analysis were: ACCORD (Action to Control Cardiovascular Risk in Diabetes), ALLHAT-HTN (Antihypertensive and Lipid-Lowering Treatment to Prevent Heart Attack), ALLHAT-LLT (Lipid-Lowering Therapy), and WHI (Women’s Health Initiative). Each trial enrolled a broad population of adult patients without prevalent cardiovascular disease. Details of the trials have been reported previously and are summarized in Supplementary Table 1.

### CPM-database Matching Process

In order to identify appropriate validation cohorts for a given CPM, we employed a hierarchical matching procedure. First, each CPM was compared with each database by non-clinical research staff to identify pairs that had grossly similar inclusion criteria and outcomes, which were then reviewed for appropriateness by clinical experts. Potential pairs passing these screening steps were carefully reviewed at a granular level, and only pairs where sufficient patient-level data existed in the trial database such that the CPM could be used to generate a predicted outcome probability for each patient were included in the analysis. Observed outcomes in the patient-level data were defined using the CPM outcome definition and prediction time horizon.

### Measuring CPM performance

The performance of CPMs in external cohorts was evaluated with measures of discrimination, calibration, and net benefit when applied to external validation cohorts. For all model validations, observed outcome events that occurred after the prediction time horizon were censored. For time-to-event models, the Kaplan-Meier estimator was used for right-censored follow up times. For binary outcome models, unobserved outcomes (ie due to loss-to-follow up prior to the prediction time horizon) were considered missing and excluded from analyses. For each CPM-database pair, the linear predictor was calculated for each patient in the database using the intercept and coefficients from the published CPM. Model discrimination was assessed using the c-statistic. The percent change in a CPM’s discrimination from the derivation cohort to the validation cohort was calculated as [(Validation c-statistic - 0.5) – (Derivation c-statistic - 0.5)] / (Derivation c-statistic - 0.5) * 100.^13^ 44 models were excluded from the assessment of decrement in c-statistic relative to derivation because the c-statistic at model development was not reported. Change in discrimination was also compared relative to the model-based c-statistic (MB-c). The MB-c is the c-statistic that would be obtained in the validation database under the assumption that the CPM is perfectly valid in the validation database.^14^ Thus, any difference between the derivation c-statistic and the validation MB-c reflects differences in case-mix, while the difference between the validation MB-c and the c-statistic in the validation cohort reflects model invalidity. Because calculation of the MB-c depends entirely on the validation cohort, MB-c could be calculated for all pairs.

Model calibration was assessed by converting the linear predictor to a predicted probability (including a specified time point if Cox proportional hazards modeling was used). From the predicted probabilities, calibration slope and Harrel’s E_AVG_ and E_90_ statistics were calculated. Harrel’s E_AVG_ and E_90_ statistics measure the mean and 90^th^ percentile, respectively, of the absolute difference between the predicted and observed event probabilities, where observed probabilities are estimated nonparametrically using locally weighted scatterplot smoothing (LOESS) curves. For this analysis E_AVG_ and E_90_ values were standardized by dividing by the outcome rate in the validation cohort to improve comparability between CPM-validation pairs. If point estimates of outcome incidences at similar time points in the CPM derivation cohort and paired validation cohort were not able to be calculated with published information, that pair was excluded from analysis of calibration.

Finally, decision curve analysis^15^ was used to estimate the net benefit of each model in each paired validation database at three distinct decision thresholds: half the outcome incidence, the outcome incidence, and double the outcome incidence. Decision curve analysis presents a comprehensive assessment of the potential population-level clinical consequences of using CPMs to inform treatment decisions by examining misclassification of patients across the full range of thresholds, while weighting the relative utility of false-positive and false-negative predictions as implicitly determined by the threshold. Models were assessed for whether they resulted in a net benefit above or below the best default strategy (treat all or treat none) at each decision threshold. Models where net benefit was equivalent to the best default strategy were considered neutral.

### Stratification by Relatedness

To explore sources of variability in model performance in external validation, we categorized each CPM-database pair based on the “relatedness” of the underlying study populations. Study populations were reviewed in detail by clinical experts on the basis of key clinical characteristics, such as inclusion/exclusion criteria, patient demographics, outcome, enrollment period, and follow-up duration. Pairs were categorized as “related” when there were no clinically relevant differences in inclusion criteria, exclusion criteria, recruitment setting, and baseline clinical characteristics. Any matches with clinically relevant differences in any criterion were categorized as “distantly related” (Supplementary Tables 2 and 3). Clinical experts scoring relatedness were blinded to the derivation c-statistic of the CPM and outcome rates in the derivation and validation cohorts.

### Model updating

We assessed the impact of model updating on discrimination, calibration, and net benefit. Models were updated using data from each paired validation database in a sequential fashion: 1) by updating the model intercept using the observed outcome rate in the validation cohort (recalibration-in-the-large); 2) by updating the intercept and rescaling all the model coefficients by the calibration slope; and 3) by re-estimating all regression coefficients using data from the validation database (but maintaining the predictors from the original model).^16^

### Statistical Analysis

Continuous variables were summarized as median [interquartile range] and proportions were summarized as N (%). Differences in various model performance measures were assessed using the Wilcoxon rank-sum test. All analyses were performed in R version 3.5.3 (R foundation for statistical computing, Vienna, Austria).

## Results

### CPM-validation cohort matching

From a set of 195 potential CPMs, 157 (80.5%) were screened as potential matches and underwent granular review to assess for sufficient patient level variable and outcome data within the 4 publicly available clinical trial databases. We matched 63 (32%) CPMs to at least one database, yielding 88 CPMs-database pairs (Figure 1). Details about the CPMs used in this analysis are summarized in Supplementary Table 4. The WHI and ACCORD databases were matched with the most CPMs (35 and 32, respectively) while ALLHAT-LLT and ALLHAT-HTN could be matched with fewer CPMs (15 and 6 CPMs, respectively).

**Figure 1:**
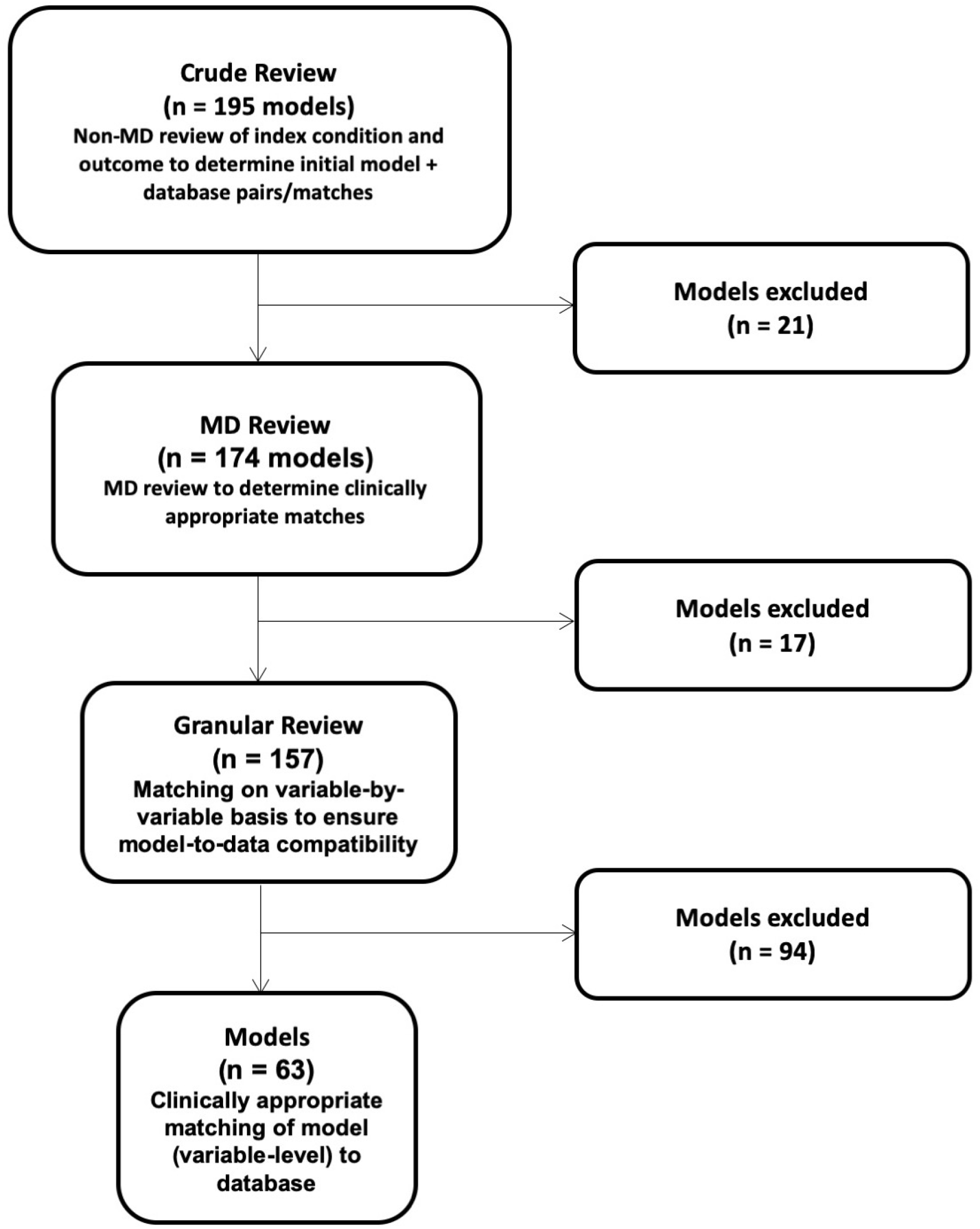
Flowchart of clinical prediction model-database matching process.

### CPM Discrimination in Independent External Validations

Of the 88 total CPM-database pairs, there were 54 pairs in which the CPM reported a c-statistic at model development. Among these, the median c-statistic in the derivation cohorts was 0.77 (IQR 0.72–0.78) and the median c-statistic at model validation was 0.63 (IQR 0.58–0.66, p <0.001 vs derivation, Table 1). Discriminative ability decreased by a median of 60% (IQR 42– 73%, Figure 2). Approximately half the loss in discriminatory power was attributable to a decrease in case-mix heterogeneity, while half was attributable to model invalidity. When stratified by relatedness, 37 (42%) pairs were graded as “related” and 51 (58%) were graded as “distantly related.” CPM-trial pairs that were related had significantly higher MB-c and validation c-statistics than pairs that were distantly related (Table 1). Median percentage decrement in discrimination among related pairs was 42% (IQR 23–46%), of which approximately two-thirds was due to a decrease in case-mix heterogeneity and one-third due to model invalidity (Figure 2). In contrast, CPM-trial pairs that were distantly related had a median percentage decrement in discrimination of 67% (IQR 60–80%, p < 0.001 vs related pairs), approximately half of which was due to case-mix heterogeneity and half due to model invalidity (Figure 2).

**Table 1.** Discriminative ability of clinical prediction models in development and validation cohorts stratified by cohort relatedness. Values are presented as median (IQR). MB-c, model-based c-statistic.

|  |  | Cohort relatedness |  |  |
| --- | --- | --- | --- | --- |
|  | All pairs<br>N=88 | Related<br>N=37 | Distantly related<br>N=51 | p-value |
| Development c-statistic* | 0.77 (0.72–0.78) | 0.77 (0.72–0.79) | 0.77 (0.72–0.78) | 0.93 |
| Validation MB-c | 0.67 (0.65–0.70)<sup>†</sup> | 0.69 (0.67–0.75)<sup>†</sup> | 0.67 (0.65–0.68)<sup>†</sup> | <0.001 |
| Validation c-statistic | 0.63 (0.58–0.66)<sup>†‡</sup> | 0.67 (0.64–0.70)<sup>†‡</sup> | 0.60 (0.56–0.62)<sup>†‡</sup> | <0.001 |

\* 34 models did not report c-statistic. Sample size for development c-statistic is 54 (18 related, 36 distantly related).
<sup>†</sup>p < 0.05 vs development
<sup>‡</sup>p < 0.05 vs MB-c

**Figure 2:**
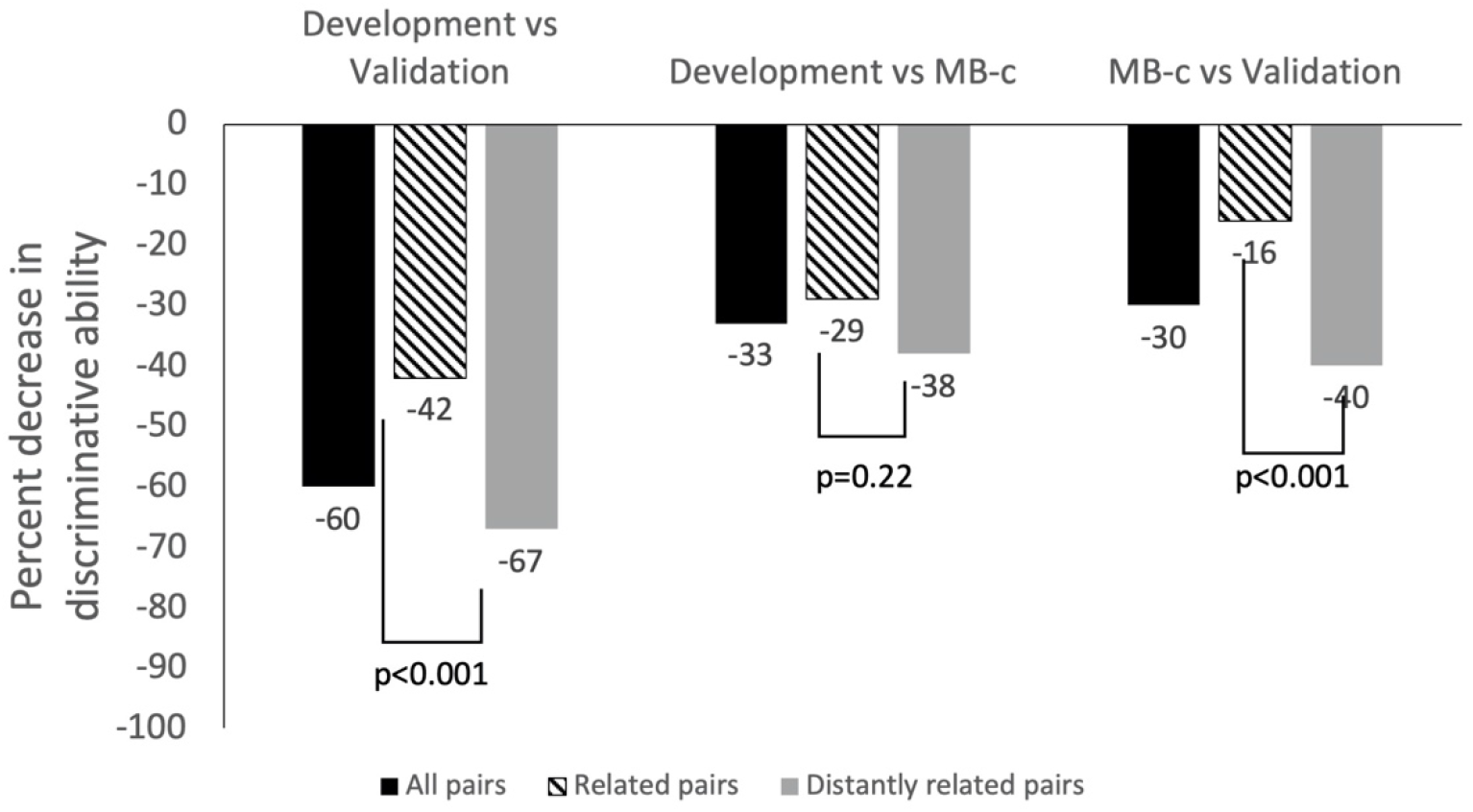
Median percent change in c-statistic in external validation cohorts. Data are presented as medians and are stratified by relatedness. MB-c, model-based c-statistic.

### CPM Calibration in Independent External Validations

The median calibration slope in the external validations was 0.62 (IQR 0.50–0.77). Median calibration slope among related pairs was 0.69 (IQR 0.59–0.84), while median calibration slope among distantly related pairs was 0.58 (IQR 0.43–0.63, p < 0.001 vs related pairs). Median E_AVG_ and median E_90_ standardized to the outcome incidence among all pairs was 0.60 (IQR 0.40–0.70) and 1.0 (IQR 0.7–1.2), respectively, and did not differ significantly between related and distantly related pairs (Table 2).

**Table 2.** Calibration performance of clinical prediction models on external validation stratified by cohort relatedness. Calibration error was measured using Harrel’s E_AVG_ and E_90_ statistics, standardized to the outcome incidence. For example, if the outcome incidence in a validation population was 5% and E_AVG_ was 0.05, standardized E_AVG_=1.0. Values are presented as median (IQR).

|  |  | Cohort relatedness |  |  |
| --- | --- | --- | --- | --- |
|  | All pairs<br>N=62 | Related<br>N=37 | Distantly related<br>N=25 | p-value |
| Calibration slope | 0.62 (0.50–0.77) | 0.69 (0.59–0.84) | 0.58 (0.43–0.63) | <0.001 |
| Standardized<br>$E_{AVG}$ | 0.60 (0.40–0.70) | 0.60 (0.50–0.70) | 0.50 (0.40–0.80) | 0.68 |
| Standardized $E_{90}$ | 1.0 (0.7–1.2) | 0.8 (0.70–1.2) | 1.1 (1.0–1.3) | 0.14 |

### Net Benefit

At a decision threshold of half the outcome incidence, 41 of 62 (66%) evaluable CPM-database pairs resulted in net benefit below the default strategy of treating all patients when applied to external validation cohorts (that is, the population-level impact of using the CPM to target treatment according to predicted risk was less favorable than that of simply treating all patients), while only 20 (32%) were beneficial relative to the default strategy (Table 3). At a threshold of twice the prevalence, 31 (50%) resulted in net benefit below the default strategy of treating no patients, and only 12 (19%) were beneficial relative to the default strategy. At a threshold equal to the outcome incidence, 5 of 62 (8%) of CPMs resulted in decreased net benefit compared with the default strategy and 51 (82%) resulted in increased net benefit.

**Table 3.**
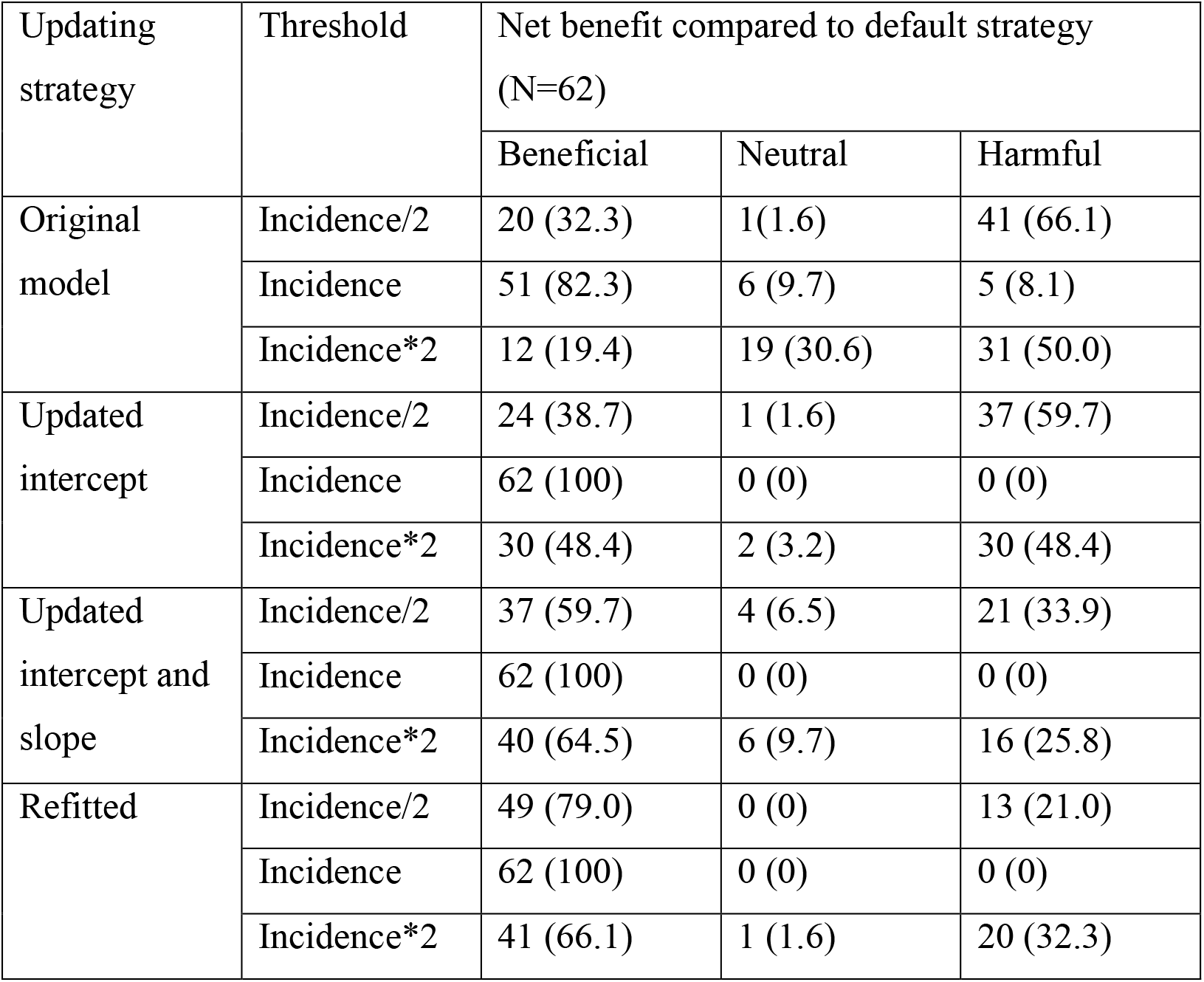
Effect of model updating on net clinical benefit in validation populations. Net benefit associated with model use before and after sequential updating procedures was assessed at 3 thresholds: incidence, half the incidence, and twice the incidence. Net benefit was considered neutral if it was within 5% of the default strategy. 26 pairs were not able to be assessed because the model predicted outcomes at a different time point than that which was used to ascertain outcomes in the clinical trial database. Values are presented as N (%).

| Updating strategy | Threshold | Net benefit compared to default strategy (N=62) |  |  |
| --- | --- | --- | --- | --- |
|  |  | Beneficial | Neutral | Harmful |
| Original model | Incidence/2 | 20 (32.3) | 1(1.6) | 41 (66.1) |
|  | Incidence | 51 (82.3) | 6 (9.7) | 5 (8.1) |
|  | Incidence*2 | 12 (19.4) | 19 (30.6) | 31 (50.0) |
| Updated intercept | Incidence/2 | 24 (38.7) | 1 (1.6) | 37 (59.7) |
|  | Incidence | 62 (100) | 0 (0) | 0 (0) |
|  | Incidence*2 | 30 (48.4) | 2 (3.2) | 30 (48.4) |
| Updated intercept and slope | Incidence/2 | 37 (59.7) | 4 (6.5) | 21 (33.9) |
|  | Incidence | 62 (100) | 0 (0) | 0 (0) |
|  | Incidence*2 | 40 (64.5) | 6 (9.7) | 16 (25.8) |
| Refitted | Incidence/2 | 49 (79.0) | 0 (0) | 13 (21.0) |
|  | Incidence | 62 (100) | 0 (0) | 0 (0) |
|  | Incidence*2 | 41 (66.1) | 1 (1.6) | 20 (32.3) |

### Effects of Updating

E_AVG_ improved by a median of 57% (IQR 43–78%) across all the CPM-trial pairs with updating of the intercept and by a median of 97% (IQR 88–100%) after updating the intercept and slope. Similar results were seen for E_90_. No further improvement in calibration error was seen with re-estimation. While updating the intercept alone eliminated the likelihood of harm relative to the default strategy at a decision threshold equivalent to the outcome incidence, this did little to reduce the likelihood of harm at more extreme decision thresholds (Table 3). At decision thresholds of half or twice the outcome incidence, likelihood of harm remained above 20% even after complete re-estimation of the model coefficients using patient-level data from the clinical trial populations.

## Discussion

Validation of CPMs is important to understanding how well they perform in new populations. In this study, we found that the vast majority of CPMs was impossible to validate on publicly available patient-level trial databases. Among the CPMs that we were able to validate, we found that discrimination and calibration deteriorated substantially when compared with the derivation cohorts. The decrease in discrimination was due to both narrower case mix in the validation cohorts as well as model invalidity, and larger decrements were observed when validation datasets were judged to be distantly related to the derivation dataset. Finally, we found that the use of CPMs on these validation cohorts without efforts at model updating may result in harmful decision making in a significant number of cases.

Of 195 potential CPMs, we were only able to evaluate 32% using patient-level data from at least one of 4 large cardiovascular RCTs. We observed two main reasons for this difficulty. First, many CPMs included predictors that were not available in the RCT databases. Secondly, some CPMs were unable to be validated because the reported methods were insufficient to recapitulate the model-building strategy in the external validation database. Researchers seeking to develop CPMs should be cognizant of both these potential issues. Models meant for broad use should prioritize the inclusion of routinely available clinical data to enable robust external validation. Additionally, transparently reporting the CPM development process can help increase trust in a model and potentially increase its uptake. To this end, the Transparent Reporting of a Multivariable Prediction Model for Individual Prognosis or Diagnosis (TRIPOD) statement outlines best practices for CPM reporting.^17^

We observed reduction in the c-statistic of the CPMs from 0.77 to 0.63 upon external validation. Similar reductions in discrimination have been reported previously for other diseases.^18–20^ Poor discriminative ability may be attributable to model invalidity (biased estimates of regression coefficients) or to a less heterogeneous patient case-mix (identifying high and low risk individuals is more difficult if the population is more homogenous). The MB-c is a useful tool to distinguish these sources because it estimates the discriminative ability of the model in a new cohort based solely on the case-mix in that cohort. The MB-c answers the question: what would the c statistic be if only the risk distribution of the validation cohort is different than in the derivation cohort, assuming the model coefficients are correct. Consequently, the difference between the c-statistic and the MB-c in external validation represents the impact of model invalidity. We found that for CPMs predicting cardiovascular events in a primary-prevention population, approximately half the decrement in discriminative ability was due to a narrower case-mix, and half was due to model invalidity. The significant amount of model invalidity we observed implies that the value of using these CPMs to guide clinical decisions in new, unselected populations may be lower than would be expected from the originally reported model performance.

Our data showed a larger decrement in discrimination when externally validating a CPM on a distantly related cohort than if the cohort were more closely related. Furthermore, the proportion of decrement in model discrimination attributable to model invalidity was higher when the cohorts were distantly related. Relatedness determinations often hinged on subtle but clinically relevant differences between cohorts, such as years of enrollment or the distribution of baseline comorbidities, that required expert clinician review to identify. However, given the especially poor discriminative ability we observed in distantly related cohorts, clinicians should carefully consider the relatedness of their population to the derivation population (with respect to important clinical characteristics such as inclusion/exclusion criteria, population demographics and comorbidities, components of a composite outcome, and follow-up time) when seeking to use an existing CPM.

Similar to the loss of discrimination, we observed a significant amount of miscalibration at external validation. The impact of both discrimination and calibration on the utility of a model can be summarized by using decision curve analysis to quantify the net benefit of using a model to guide clinical decision making at various decision thresholds. Our analysis showed that across all CPM-validation pairs, there was a high likelihood that use of the CPM in that validation population would result in harm at the population level relative to a default strategy of treating all or treating no patients, particularly when the decision threshold was distant from the overall outcome incidence. At these thresholds, harm relative to the best default strategy was seen in more than half of model-cohort pairs. We found that sequential model updating procedures could reduce the likelihood of harm. Updating the intercept alone (recalibration-in-the-large) eliminated the potential for harm at a decision threshold equal to the outcome incidence, but did little to reduce the likelihood of harm at the extreme decision thresholds we evaluated. More intensive model updating procedures did reduce the likelihood of harm at extreme decision thresholds, but this likelihood remained > 20% even with complete re-estimation of model coefficients. These findings highlight that applying a published CPM to a new population without updating requires great caution because there is a high likelihood that the model could worsen, rather than improve decision making, due to systematic misestimation of risk. Selecting the most appropriate updating procedure will likely depend on the clinical scenario, the size of the validation dataset, and the difference between the overall outcome incidence and the decision threshold of interest.^16^

Concerns about harm from the use of poorly calibrated models garnered substantial attention when the 2013 American College of Cardiology/American Heart Association (ACC/AHA) Cholesterol Treatment Guidelines, which recommended primary prevention statin therapy for nondiabetic individuals with an estimated 10-year CVD risk of ≥ 7.5%, were released.^9^ CVD risk is estimated from the PCE, which were derived from multiple population cohorts studied in the 1990s and showed good discrimination (c-statistics 0.7–0.8) across multiple ethnic groups.^21^ However, when the PCE were applied to contemporary cohorts, CVD risk was overestimated by 75–150%, indicative of significant miscalibration.^10^ This suggests that among the 30 million Americans who might qualify for statin initiation based on the 2013 guidelines, the true 10-year CVD risk in the lowest risk individuals may actually fall well below the 7.5% threshold, resulting in potentially harmful over-treatment. This has led some to argue for “trial-based” guidelines, where treatment recommendations are applied uniformly to populations defined by RCT inclusion criteria, rather than “risk-based” guidelines.^10^ While our work and that of others has pointed to the potential advantages of risk-based decision making,^6,22^ the results of the present analysis underscore the dangers associated with poor calibration and suggest that the advantages of risk-based decision making might only be fully realized when models are frequently updated on the populations to which they are applied.^23^

### Limitations

There are several limitations to this analysis. We validated CPMs in cohorts from clinical trials, rather than real-world cohorts, which are expected to be more homogenous than real-world cohorts. However, our use of the MB-c enabled us to disentangle the decrement in discrimination attributable to the narrower case mix in the clinical trial cohort from that attributable to invalid model coefficients. The high proportion of decrement in discrimination attributable to model invalidity seen in this analysis implies significant decrements in model performance would be expected even in more heterogenous validation cohorts. The available databases used for external validation represent patients across different time periods, and differences in primary prevention treatment may have changed the relationships between predictors and outcomes between the development and validation cohorts evaluated here. Contemporaneous validation cohorts may have shown smaller decrements in discrimination or validation, though such a finding would underscore our conclusion about the importance of considering the degree of similarity to the derivation cohort when applying a CPM. Each CPM was only able to be externally validated a small number of times. A given CPM may perform differently when validated against different cohorts, and more research is required to understand the sources of this variation before validation performance can be used to grade the quality of a model. Our relatedness categorization was one such attempt, but requires content area expertise, is inherently subjective, and is difficult to generalize to CPMs for other clinical domains. Finally, our net benefit analyses focused on three arbitrary decision thresholds and were not informed by the relative cost of over-treatment versus under-treatment in the specific clinical context.

## Conclusions

This analysis highlights several important aspects regarding the landscape of CPMs for primary prevention of cardiovascular disease. Many models are difficult to externally validate because they are not reported transparently or because they include specialized measurements not readily available during routine care. Among those that we were able to validate, significant decrements in discrimination and severe miscalibration were noted, resulting in potentially harmful decision making if used on new cohorts. While some loss of discrimination was due to the narrower case-mix of the validation populations, half of the decrement was attributable to model invalidity. More effort to externally validate existing CPMs is warranted to better understand CPM performance in new populations. Appling existing CPMs to new populations without model updating should be done with caution, as the likelihood of miscalibration is high, and systematic over- or under-estimation of risk can lead to harmful decision making manifested by systematically over- or under-treating patients.

## Supporting information

PRISMA checklist

## Data Availability

The Tufts Predictive Analytics and Comparative Effectiveness (PACE) CPM Registry

http://pacecpmregistry.org/

