## Supplemental Table 1 for "Clinical Prediction Models for Primary Prevention of Cardiovascular Disease: Validity in Independent Cohorts"

**Word count:** TBD

**Short title:** Validation of Primary Prevention Cardiovascular Prediction Models

**Keywords:** clinical prediction model, validation, regression, primary prevention

**Address for Correspondence:**

David M. Kent, MD, MS

**Supplementary Table 1. Summary of clinical trial databases used as external validation cohorts.** Cr, creatinine; CHD, coronary heart disease; CVD, cardiovascular disease; DM, diabetes mellitus; HR, hazard ratio; HRT, hormone replacement therapy; HTN, hypertension; hx, history; LDL, low-density lipoprotein; MI, myocardial infarction; RR, risk ratio; US, united states

| Trial | Sample size | Enrollment Period | Locations | Target population | Selected baseline characteristics | Selected baseline medications | Intervention | Outcome | Effect size | Outcome incidence | Mean length of follow up |
| --- | --- | --- | --- | --- | --- | --- | --- | --- | --- | --- | --- |
| ACCORD | 10251 | 2001–2005 | 77 sites in US and Canada | Age 40–79, type 2 DM with HbA1C > 7.5%, Cr < 1.5 mg/dL | Mean age 62.2, mean BMI 32.3 kg/m^2^, median HbA1c 8.1%, 38% female, 59% smoking hx, 35% CVD hx | insulin: 35%  metformin: 60%  any antihypertensive: 85% | Intensive therapy (target HbA1c < 6%) vs standard therapy (target HbA1C 7–7.9%) | Composite of CV death, nonfatal MI, or nonfatal stroke | Intensive vs standard therapy HR 0.9 (95% CI 0.78–1.04) | Intensive therapy: 6.9%  Standard therapy: 7.2% | 3.5 years |
| ALLHAT-HTN | 33357 | 1994–1998 | 623 sites in North America | Age ≥ 55, HTN, CVD risk | Mean age 67, mean BMI 30 kg/m^2^, 47% female, 36% DM, 22% current smoker, 52% CVD hx | aspirin: 36% | chlorthalidone vs amlodipine vs lisinopril | Composite of CHD death or nonfatal MI | Amlodipine vs chlorthalidone RR 0.98 (95% CI 0.9–1.7); lisinopril vs chlorthalidone RR 0.99 (95% CI 0.91–1.08) | Chlorthalidone: 8.9%  Amlodipine: 8.8%  Lisinopril: 8.8% | 4.9 years |
| ALLHAT-LLT | 10355 | 1994–1998 | 513 sites in North America | Age ≥ 55, HTN, LDL 120–189 mg/dL | Mean age 66, mean BMI 30 kg/m^2^, 49% female, 35% DM, 23% current smoker, 14% CVD hx | aspirin: 31% | pravastatin vs usual care | All-cause mortality | Pravastatin vs usual care RR 0.99 (95% CI 0.89–1.11) | Pravastatin: 14.9%  Usual care: 15.3% | 4.8 years |
| WHI | 16608 | 1993–1998 | 40 sites in US | Women age 50–79 | Mean age 63, mean BMI 28.5 kg/m^2^, 4.5% DM, 50% smoking hx, 3% angina hx | aspirin: 20%  statin: 7% | HRT vs placebo | Composite of CHD death or nonfatal MI | HRT vs placebo HR 1.29 (95% CI 1.02–1.63) | HRT: 0.37%  Placebo: 0.3% | 5.2 years |
