## Supplemental Table 2 for "Clinical Prediction Models for Primary Prevention of Cardiovascular Disease: Validity in Independent Cohorts"

**Short title:** Validation of Primary Prevention Cardiovascular Prediction Models

**Keywords:** clinical prediction model, validation, regression, primary prevention

**Address for Correspondence:**

David M. Kent, MD, MS

**Supplementary Table 2. Example of “related” CPM-clinical trial cohorts.** Cohorts were compared on the factors in the table and scored as “related” or “distantly related” as described in the manuscript. For this pair, no clinically relevant differences were found, and the pair was scored as “related.” CPM, clinical prediction model; Cr, creatinine; DM, diabetes mellitus; HbA1c, hemoglobin A1c; NR, not reported

|  | CPM cohort | Clinical trial cohort |
| --- | --- | --- |
| Recruitment details |  |  |
| Inclusion criteria | All newly referred diabetic patients to the Prince of Wales Hospital in Hong Kong | Type II DM; HbA1c ≥ 7.5%; age 40-79 years and cardiovascular disease OR age 55-79 with significant atherosclerosis, albuminuria, left ventricular hypertrophy, or at least two additional risk factors for cardiovascular disease (dyslipidemia, hypertension, current status as a smoker, or obesity) |
| Exclusion criteria | Patients with type 1 diabetes | Frequent or recent serious hypoglycemic events, unwillingness to do home glucose monitoring or inject insulin, BMI > 45, Cr > 1.5 mg/dL, or other serious illness |
| Recruitment setting | Outpatient | Community |
| Enrollment period | 1995-2005 | 2001-2005 |
| Risk factors |  |  |
| Diabetes | 100% | 100% |
| HTN | 35% | NR |
| HLD | NR | NR |
| Outcome details |  |  |
| Outcome | stroke | stroke |
| Duration of follow up | 5 years | 3.5 years |
