## Supplemental Table 3 for "Clinical Prediction Models for Primary Prevention of Cardiovascular Disease: Validity in Independent Cohorts"

**Short title:** Validation of Primary Prevention Cardiovascular Prediction Models

**Keywords:** clinical prediction model, validation, regression, primary prevention

**Address for Correspondence:**

David M. Kent, MD, MS

**Supplementary Table 3. Example of “distantly related” CPM-clinical trial cohorts.** The cohorts were distantly related because of clinically significant differences in inclusion criteria (CPM cohort excluded patients with prevalent coronary disease while the clinical trial cohort included such patients) and the proportion of diabetics. CHD, coronary heart disease; CPM, clinical prediction model; Cr, creatinine; DM, diabetes mellitus; HbA1c; hemoglobin A1c; MI, myocardial infarction; NR, not reported

|  | CPM cohort | Clinical trial cohort |
| --- | --- | --- |
| Recruitment details |  |  |
| Inclusion criteria | Black and Non-Black men and women, age 45–64 at entry. | Type II DM; HbA1c ≥ 7.5%; age 40-79 years and cardiovascular disease OR age 55-79 with significant atherosclerosis, albuminuria, left ventricular hypertrophy, or at least two additional risk factors for cardiovascular disease (dyslipidemia, hypertension, current status as a smoker, or obesity) |
| Exclusion criteria | Prevalent CHD or CHD identified at the baseline visit | Frequent or recent serious hypoglycemic events, unwillingness to do home glucose monitoring or inject insulin, BMI > 45, Cr > 1.5 mg/dL, or other serious illness |
| Recruitment setting | Community | Community |
| Enrollment period | 1987-1989 | 2001-2005 |
| Risk factors |  |  |
| Diabetes | 7% | 100% |
| HTN | 35% | NR |
| HLD | 20% | NR |
| Outcome details |  |  |
| Outcome | CHD (MI or fatal CHD) | Nonfatal MI or death from cardiovascular causes |
| Duration of follow up | 10 years | 3.5 years |
