## Supplemental Table 4 for "Clinical Prediction Models for Primary Prevention of Cardiovascular Disease: Validity in Independent Cohorts"

**Short title:** Validation of Primary Prevention Cardiovascular Prediction Models

**Keywords:** clinical prediction model, validation, regression, primary prevention

**Address for Correspondence:**

David M. Kent, MD, MS

**Supplementary Table 4. Summary of clinical prediction models externally validated against clinical trial databases.** ACR, albumin to creatinine ratio; AF, atrial fibrillation; ASCVD, atherosclerotic cardiovascular and cerebrovascular disease; BMI, body mass index; CABG, coronary artery bypass graft surgery; CeVD, cerebrovascular disease; CHD, coronary heart disease; CRP, C-reactive protein; CVD, cardiovascular disease; DBP, diastolic blood pressure; EKG, electrocardiogram; HbA1c, hemoglobin A1c; HDL, high-density lipoprotein; HLD, hyperlipidemia; HTN, hypertension; IMT, intimal-medial thickness; LDL, low-density lipoprotein; LVH, left ventricular hypertrophy; MI, myocardial infarction; NR, not reported; PCI, percutaneous coronary intervention; PMID, PubMed ID; SBP, systolic blood pressure; TIA, transient ischemic attack; WBC, white blood count

| PMID | Population | Outcome | Model sample size | Years of enrollment | Mean or median follow up time (years) | Number of events | Mean age or age range | Male (%) | Diabetes (%) | Current smoker (%) | Variables in final model |
| --- | --- | --- | --- | --- | --- | --- | --- | --- | --- | --- | --- |
| 1985385 | Members of the Framingham Heart Study and Framingham Offspring Study cohorts, without baseline CVD or cancer (other than basal cell carcinomas). | 10-Year CHD (MI and CHD death plus angina pectoris and coronary insufficiency) | 5573 | 1968 - 1975 | 12 | NR | 30-74 | 46.5% | 6.0% | 39.7% | Gender, age, SBP, smoking, total cholesterol, HDL, diabetes, LVH on EKG |
| 1985385 | Members of the Framingham Heart Study and Framingham Offspring Study cohorts, without baseline CVD or cancer (other than basal cell carcinomas). | 10-Year MI (including silent and unrecognized) | 5573 | 1968 - 1975 | 12 | NR | 30-75 | 46.5% | 6.0% | 39.7% | Gender, age, SBP, smoking, total cholesterol, HDL, diabetes, LVH on EKG |
| 1985385 | Members of the Framingham Heart Study and Framingham Offspring Study cohorts, without baseline CVD or cancer (other than basal cell carcinomas). | 10-Year CHD Mortality (including sudden and nonsudden) | 5573 | 1968 - 1975 | 12 | NR | 30-74 | 46.5% | 6.0% | 39.7% | Gender, age, SBP, smoking, total cholesterol, HDL, diabetes, LVH on EKG |
| 1985385 | Members of the Framingham Heart Study and Framingham Offspring Study cohorts, without baseline CVD or cancer (other than basal cell carcinomas). | 10-Year Stroke (including transient ischemic attack) | 5573 | 1968 - 1975 | 12 | NR | 30-74 | 46.5% | 6.0% | 39.7% | Gender, age, SBP, smoking, total cholesterol, HDL, diabetes, LVH on EKG |
| 1985385 | Members of the Framingham Heart Study and Framingham Offspring Study cohorts, without baseline CVD or cancer (other than basal cell carcinomas). | 10-Year CVD (MI, CHD death plus angina pectoris and coronary insufficiency, congestive heart failure, and peripheral vascular disease) | 5573 | 1968 - 1975 | 12 | NR | 30-74 | 46.5% | 6.0% | 39.7% | Gender, age, SBP, smoking, total cholesterol, HDL, diabetes, LVH on EKG |
| 1985385 | Members of the Framingham Heart Study and Framingham Offspring Study cohorts, without baseline CVD or cancer (other than basal cell carcinomas). | 10-Year CHD Mortality (including sudden and nonsudden) | 5573 | 1968 - 1975 | 12 | NR | 30-74 | 46.5% | 6.0% | 39.7% | Gender, age, SBP, smoking, total cholesterol, HDL, diabetes, LVH on EKG |
| 1985385 | Members of the Framingham Heart Study and Framingham Offspring Study cohorts, without baseline CVD or cancer (other than basal cell carcinomas). | 10-Year CHD (MI and CHD death plus angina pectoris and coronary insufficiency) | 5573 | 1968 - 1975 | 12 | NR | 30-74 | 46.5% | 6.0% | 39.7% | Gender, age, DBP, smoking, total cholesterol, HDL, diabetes, LVH on EKG |
| 1985385 | Members of the Framingham Heart Study and Framingham Offspring Study cohorts, without baseline CVD or cancer (other than basal cell carcinomas). | 10-Year MI (including silent and unrecognized) | 5573 | 1968 - 1975 | 12 | NR | 30-74 | 46.5% | 6.0% | 39.7% | Gender, age, DBP, smoking, total cholesterol, HDL, diabetes, LVH on EKG |
| 1985385 | Members of the Framingham Heart Study and Framingham Offspring Study cohorts, without baseline CVD or cancer (other than basal cell carcinomas). | 10-Year CHD Mortality | 5573 | 1968 - 1975 | 12 | NR | 30-74 | 46.5% | 6.0% | 39.7% | Gender, age, DBP, smoking, total cholesterol, HDL, diabetes, LVH on EKG |
| 1985385 | Members of the Framingham Heart Study and Framingham Offspring Study cohorts, without baseline CVD or cancer (other than basal cell carcinomas). | 10-Year Stroke (including transient ischemic attack) | 5573 | 1968 - 1975 | 12 | NR | 30-74 | 46.5% | 6.0% | 39.7% | Gender, age, DBP, smoking, total cholesterol, HDL, diabetes, LVH on EKG |
| 1985385 | Members of the Framingham Heart Study and Framingham Offspring Study cohorts, without baseline CVD or cancer (other than basal cell carcinomas). | 10-Year CVD (MI, CHD death plus angina pectoris and coronary insufficiency, congestive heart failure, and peripheral vascular disease) | 5573 | 1968 - 1975 | 12 | NR | 30-74 | 46.5% | 6.0% | 39.7% | Gender, age, DBP, smoking, total cholesterol, HDL, diabetes, LVH on EKG |
| 1985385 | Members of the Framingham Heart Study and Framingham Offspring Study cohorts, without baseline CVD or cancer (other than basal cell carcinomas). | 10-Year CVD Mortality | 5573 | 1968 - 1975 | 12 | NR | 30-74 | 46.5% | 6.0% | 39.7% | Gender, age, DBP, smoking, total cholesterol, HDL, diabetes, LVH on EKG |
| 2003301 | Men and Women from the Framingham Study Cohort | 10-Year incident ischemic stroke | 5734 | 1968 - 1975 | 10 | 472 | 55-84 | 41.4% | 9.0% | 29.5% | Age, SBP, Antihypertensive therapy, Diabetes melllitus, smoking, CVD, AF, LVH |
| 2003301 | Women from the Framingham Study Cohort | 10-Year incident ischemic stroke | 3362 | 1968 - 1975 | 10 | 259 | 65.8 | 0 | 7.9% | 26.4% | Age, SBP, Antihypertensive therapy, Diabetes melllitus, smoking, CVD, AF, LVH |
| 8266381 | Framingheart participants without prior stroke | 10-Year incident stroke (brain infarct, TIA, cerebral embolus, intracranial hemorrhage) | 5734 | 1968 - 1975 | 10 | 472 | 55-84 | 46.5% | 6.0% | 39.7% | Age, SBP, Antihypertensive therapy, Diabetes melllitus, smoking, CVD, AF, LVH |
| 8266381 | Female Framingheart participants without prior stroke | 10-Year incident stroke (brain infarct, TIA, cerebral embolus, intracranial hemorrhage) | 3362 | 1968 - 1975 | 10 | 259 | 66.1 | 0.0% | 7.9% | 26.4% | Age, SBP, Antihypertensive therapy, Diabetes melllitus, smoking, CVD, AF, LVH |
| 9425461 | Residents of the town of Busselton in Western Australia | 10-Year CHD mortality | 3891 | 1966 - 1981 | NR | 187 | 40-74 | 49.4% | NR | 34.4% | Age, SBP, Cholesterol, Smoking |
| 9425461 | Women in the town of Busselton in Western Australia | 10-Year CHD mortality | 1968 | 1966 - 1981 | NR | 71 | 55.8 | 0.0% | NR | 24.0% | Age, SBP, Cholesterol, Smoking |
| 9425461 | Residents of the town of Busselton in Western Australia | 10-Year CHD mortality | 3891 | 1966 - 1981 | NR | 187 | 40-74 | 49.4% | NR | 34.4% | Age, SBP, Cholesterol, Smoking, BMI |
| 9425461 | Women in the town of Busselton in Western Australia | 10-Year CHD mortality | 1968 | 1966 - 1981 | NR | 71 | 55.8 | 0.0% | NR | 24.0% | Age, SBP, Cholesterol, Smoking, BMI |
| 9603539 | Men from the Framingham Heart Study Population | 10-Year incident CHD (angina pectoris, MI, coronary insufficiency, and CHD death) | 5345 | 1971 - 1974 | NR | NR | 30-74 | 46.6% | 4.5% | 39.0% | Age, Total Cholesterol, HDL, Blood Pressure, Diabetes, Smoking |
| 9603539 | Women from the Framingham Heart Study Population | 10-Year incident CHD (angina pectoris, MI, coronary insufficiency, and CHD death) | 2856 | 1971 - 1974 | NR | NR | 49.8 | 0 | 4.0% | 37.7% | Age, Total Cholesterol, HDL, Blood Pressure, Diabetes, Smoking |
| 11724655 | All newly diagnosed diabetic patients from 23 UK general practioner offices without prior MI, angina or heart failure | 20-Year incident fatal or nonfatal MI or sudden death | 4540 | 1977 - 1991 | 10.3 | NR | 52.8 | 57.1% | 100.0% | 30.2% | Age of diabetes diagnosis, sex, ethnicity, smoking, HbA1C, SBP, total cholesterol, HDL, time in years since diagnosis of diabetes |
| 14514579 | Subset of Women from the Atherosclerosis Risk in Communities (ARIC) study (subset was diabetic patients from 4 US communities without baseline CHD) | 10-Year fatal and non-fatal coronary events | 861 | 1987 - 1989 | 10.2 | 108 | 45-64 | 0.0% | 100.0% | 25.4% | Age, Race (Black vs White), Total cholesterol, HDL, SBP, HTN Medication, Smoking, BMI, Waist-to-hip ratio, Sport activity, Keys Score, Serum Creatinine, Serum albumin, WBC, Factor VIII, LVH, IMT |
| 14514579 | Subset of adults from the Atherosclerosis Risk in Communities (ARIC) study (subset was diabetic patients from 4 US communities without baseline CHD) | 10-Year fatal and non-fatal coronary events | 1500 | 1987 - 1989 | 10.2 | 257 | 55 | 42.6% | 100.0% | 27.0% | Age, Race (Black vs White), Total cholesterol, HDL, SBP, HTN Medication, Smoking, BMI, Waist-to-hip ratio, Sport activity, Keys Score, Serum Creatinine, Serum albumin, WBC, Factor VIII, LVH, IMT |
| 15173150 | Adults from 11 provinces of China without prior MI nor angina | 10-Year hard CHD events (coronary death and MI) | 30121 | 1992 - 1993 | 12 | 191 | 35-64 | 53.3% | 5.5% | 33.3% | Age, Blood Pressure, Smoking, Diabetes, Total Cholesterol, HDL |
| 15173150 | Women from 11 provinces of China without prior MI nor angina | 10-Year hard CHD events (coronary death and MI) | 14056 | 1992 - 1993 | 12 | 54 | 35-64 | 0.0% | 5.0% | 4.0% | Age, Blood Pressure, Smoking, Diabetes, Total Cholesterol, HDL |
| 15659467 | CUORE study cohorts in Italy; Italian men without coronary heart disease | 10-Year fatal and non-fatal coronary events | 6865 | 1985 - 1995 | 9.1 | 312 | 35-69 | 100.0% | NR | NR | Age, LDL , HDL, Triglycerides, SBP, Smoking, Family history of MI, Diabetes |
| 15662552 | adults from the DECODE cohort (population-based or occupational-based groups in Europe) | 5-Year Cardiovascular Death | 25413 | NR | NR | 357 | 30-74 | 65.0% | 5.0% | 40.5% | Age, Fasting Plasma Glucose, Smoking, SBP, Cholesterol |
| 15662552 | adults from the DECODE cohort (population-based or occupational-based groups in Europe) | 10-Year Cardiovascular Death | 25413 | NR | NR | 791 | 30-74 | 65.0% | 5.0% | 40.5% | Age, Fasting Plasma Glucose, Smoking, SBP, Cholesterol |
| 16085199 | Male Chinese Steelworkers | 10-Year CHD (fatal MI, nonfatal MI, angina pectoris) | 4400 | 1974 - 1980 | 13.5 | 55 | 45 | 100.0% | NR | 74.0% | Age, SBP, Total Cholesterol, BMI, Smoking |
| 16085199 | Male Chinese Steelworkers | 10-Year Ischemic Stroke | 4400 | 1974 - 1980 | 13.5 | 49 | 45 | 100.0% | NR | 74.0% | Age, SBP, Total Cholesterol, Smoking |
| 16732001 | Tayside Scotland population with Type 2 Diabetes without previous cardiovascular disease event | 5-Year MI (fatal and nonfatal) or CHD death | 4569 | 1995 - 2004 | NR | 243 | 59.5 | 52.6% | 100.0% | 23.5% | Age at Diagnosis, Duration of Diabetes, HbA1c, Smoking, Sex, SBP, Treated HTN, Total Choelsterol, Height |
| 17088464 | USA-PRC Cohort (Local residents in and around Beijing and Guangzhou, China) | 10-Year Incident Ischemic CVD (MI and Ischemic Stroke) | 9903 | 1983 - 1984 | NR | 371 | 45.5 | 49.4% | 2.0% | 44.7% | Age, Systolic Blood Pressure, BMI, Total Cholesterol, Smoking, Diabetes |
| 17088464 | Women from the USA-PRC Cohort (Local residents in and around Beijing and Guangzhou, China) | 10-Year Incident Ischemic CVD (MI and Ischemic Stroke) | 5013 | 1983 - 1984 | NR | 147 | 45 | 0.0% | 2.0% | 17.0% | Age, SBP, BMI, Total Cholesterol, Smoking, Diabetes |
| 17192335 | Type 2 Diabetic Patients in Hong Kong from a single center | 5-Year incident stroke | 7209 | 1995 - 2005 | 5.4 | 372 | 57 | 45.5% | 100.0% | 20.3% | Age, HbA1C, ACR, History of CHD |
| 17478150 | Men from the Atherosclerosis Risk in Communities (ARIC) study without prior/baseline CHD | 10 Year CHD events (MI, Fatal CHD, Cardiac Procedure) | 6239 | 1987 - 1989 | NR | NR | 54.4 | 100.0% | 6.2% | 27.5% | Age, Diabetes, HTN, HLD, Smoking, Physical Activity, Family History |
| 18036028 | Employees of 52 companies and local government | 10-Year Major Coronary Event (sudden cardiac death, fatal or non-fatal MI) | 26975 | 1978 - 1996 | 9.4 | 511 | 45.7 | 68.4% | 6.2% | 29.0% | Age, SBP LDL, HDL, Triglycerides, diabetes, smoking history, family history of CVD |
| 18036028 | Employees of 52 companies and local government | 10-Year TIA or stroke (both ischemic and hemorrhagic) | 26975 | 1978 - 1996 | 9.4 | 511 | 20-79 |  |  |  | Age, SBP LDL, HDL, Triglycerides, diabetes, smoking history, family history of CVD |
| 18212285 | Adult Framingheart participants without prior CVD | 10-Year incident CVD events (CHD, stroke, peripheral artery disease, MI) | 8491 | 1968 - 1975 | 14 | 1174 | 30-74 | 46.7% | 5.0% | 34.7% | Age, Total Cholesterol, HDL, SBP, Treatment for HTN, Smoking, Diabetes |
| 18212285 | Female Framingheart participants without prior CVD | 10-Year incident CVD events (CHD, stroke, peripheral artery disease, MI) | 4522 | 1968 - 1975 | 14 | 456 | 49.1 | 0.0% | 3.8% | 34.2% | Age, Total Cholesterol, HDL, SBP, Treatment for HTN, Smoking, Diabetes |
| 18591403 | Type 2 diabetic patients without previous CVD from the Swedish National Diabetes Register | 5-Year composite of fatal or nonfatal stroke, fatal or nonfatal MI, sudden cardiac death, unstable angina, PCI or CABG | 11646 | 1996 - 1998 | 5.6 | 990 | 18-70 | 57.0% | 100.0% | 18.0% | A1C, onset age of diabetes, diabetes duration, sex, BMI, smoking, SBP, antihypertensive or lipid reducing therapy |
| 18591432 | Framingham Offspring Study participants | First CHD event (angina pectoris, MI, cardiac death) | 4780 | 1971 | 24 | 492 | 36.7 | 48.4% | 2.8% | 45.2% | Age, Sex, Smoking, BMI, Total Cholesterol-to-HDL ratio, SBP, diabetes mellitus |
| 18591432 | Framingham Offspring Study participants | First CeVD event (ischemic stroke, transient ischemic attack, stroke-related death) | 4780 | 1971 | 24 | 111 | 36.7 | 48.4% | 2.8% | 45.2% | Age, Sex, Smoking, BMI, Total Cholesterol-to-HDL ratio, SBP |
| 18762707 | Japanese Cohort with hypercholesteremia (total cholesterol 220-270 mg/dL) and no history of ischemic heart disease or ischemic stroke, including post-menopausal women | 5-Year CHD (fatal and nonfatal MI, angina pectoris, cardiac/sudden death, angioplasty) | 7760 | 1994 - 2004 | NR | 138 | 58.3 | 31.5% | NR | NR | Age, Sex, BMI, SBP, DBP, Hypertension, Glucose abnormality, Smoking, triglycerides, LDL, HDL, total cholesterol, lipoprotein(a) |
| 18997194 | US men without diabetes, heart disease, diabetes mellitus and cancer | 10-Year incident CVD events (nonfatal MI, nonfatal stroke, coronary revascularization, or cardiovascular death) | 10724 | 1995 | 10.8 | 1294 | 63 | 100.0% | 0.0% | 3.2% | Age, Blood Pressure, Smoking, Total Cholesterol, HDL, high-sensitivity CRP, parental history of MI before age 60 years |
| 19763133 | Residents of the town of Hisayama, Japan, aged 40 and older | incident development of CVD (MI, silent MI, sudden cardiac death, need for bypass or angioplasty) within 14 years of enrollment | 1756 | 1988 | 14 | 216 | 59 | 0.4% | 11.0% | 24.0% | Age, Gender, SBP, Diabetes, LDL, HDL, Smoking |
| 20197631 | Nested case-control group of Japanese male workers enrolled from 76 workplaces by occupational physicians | MI | 612 | 1997 - 2000 | NR | 204 | 35-65 | 100.0% | 5.4% | 50.8% | Blood Pressure, LDL, HDL, triglycerides, glucose, smoking |
| 20197631 | Nested case-control group of Japanese male workers | MI | 612 | 1997 - 2000 | NR | 204 | 35-65 | 100.0% | 5.4% | 50.8% | Blood Pressure, LDL, HDL, triglycerides, glucose, smoking |
| 20197631 | Nested case-control group of Japanese male workers | MI | 612 | 1997 - 2000 | NR | 204 | 35-65 | 100.0% | 5.4% | 50.8% | Blood Pressure, LDL, HDL, triglycerides, glucose, smoking |
| 20447530 | Case-Control Subset of the Women's Health Initiative, including cases of CHD, stroke, and venous-thromboembolism with controls matched on age, randomization, date, hysterectomy status, and CVD at baseline. | 5-Year CHD (nonfatal MI, fatal MI, incident silent MI) | 1064 | 1993 - 1998 | NR | 321 | NR | 0.0% | 9.7% | 12.3% | Age, Total Cholesterol, HDL, SBP, Treatment for HTN, Smoking, Diabetes Status |
| 20671251 | Community members from town 30km north of Taipei, Taiwan without prior stroke events | 10-Year incident ischemic or hemorrhagic stroke (excluding TIA) | 3513 | 1990 | 15.9 | 240 | 55 | 47.0% | 13.0% | 36.8% | Age, Gender, SBP, DBP, family history of stroke, AF, diabetes |
| 20671251 | Community members from town 30km north of Taipei, Taiwan without prior stroke events | 10-Year incident ischemic or hemorrhagic stroke (excluding TIA) | 3513 | 1990 | 15.9 | 240 | 55 | 47.0% | 13.0% | 36.8% | Age, Gender, SBP, DBP, family history of stroke, AF, diabetes, total cholesterol, WBC, fasting glucose |
| 22374565 | patients from the Prevention of MetS and Multi-metabolic Disorders in Jiangu Province of China Study (PMMJS) | Incident CHD and stroke | 3598 | 2000 - 2004 | 6.3 | 82 | 50.2 | 40.3% | NR | NR | Age, Waist Circumference, Triglycerides, SBP, DBP, Fasting Plasma Glucose, HDL |
| 22498473 | NR | 10-Year all-cause mortality | NR | NR | NR | NR | NR | NR | NR | NR | Age, SBP, Smoking, Total Cholesterol |
| 22498473 | NR | 10-Year CVD mortality | NR | NR | NR | NR | NR | NR | NR | NR | Age, SBP, Smoking, Total Cholesterol |
| 22503568 | Community members from town 30km north of Taipei, Taiwan without prior coronary events | 10-Year incident coronary artery disease event | 3430 | 1990 | 15.9 | 171 | 55 | 48.0% | 13.0% | 36.8% | Gender, Age, BMI, SBP, Smoking |
| 22503568 | Community members from town 30km north of Taipei, Taiwan without prior coronary events | 10-Year incident coronary artery disease event | 3430 | 1990 | 15.9 | 171 | 55 | 48.0% | 13.0% | 36.8% | Gender, Age, BMI, SBP, Smoking, Total cholesterol, HDL, LDL |
| 22503568 | Community members from town 30km north of Taipei, Taiwan without prior coronary events | 10-Year incident coronary artery disease event | 3430 | 1990 | 15.9 | 171 | 55 | 48.0% | 13.0% | 36.8% | Gender, Age, BMI, SBP, Smoking, Total cholesterol, HDL, LDL, LVH, WBC |
| 23407372 | Northern European hypertensive patients without prior stroke or MI | 5-Year first cardiovascular event (cardiovascular death, MI, or stroke) | 15955 | 1998 - 2000 | 5.4 | 1240 | NR | NR | NR | NR | Age, Sex, Smoking, Diabetes, Previous HTN treatment, SBP |
| 23449266 | Japan Public Health Center study cohort II: all residents with Japanese nationality (excluding history of CVD) | 10-Year risk of stroke (hemorrhagic or ischemic) | 15672 | 1993 - 1994 | NR | 790 | 40-69 | 33.9% | 4.6% | 15.4% | Age, Sex, Smoking, BMI, Blood Pressure, Diabetes Mellitus, HTN Medication |
| 23860709 | US-based NHANES III | 10-Year fatal CVD | 15454 | 1988 - 1994 | NR | 1716 | NR | NR | NR | NR | Age, Sex, Smoking, SBP, Total Cholesterol, Total cholesterol to HDL ratio, Glucose, HbA1C |
| 24239921 | women free of atherosclerotic cardiovascular disease (ASCVD); taken from several long-standing population-based cohort studies funded by the NHLBI including ARIC, Cardiovascular Health Study, CARDIA, and Framingham) | 10-Year risk of hard ASCVD (first occurrence of nonfatal MI or CHD death, or fatal or nonfatal stroke) | 13881 | NR | NR | 1192 | 40-79 | 0 | NR | NR | Age, Total Cholesterol, HDL, SBP, Diabetes, Smoking |
| 24239921 | white men free of atherosclerotic cardiovascular disease (ASCVD) from 40 to 79 years of age; taken from several long-standing population-based cohort studies funded by the NHLBI | 10-Year risk of hard ASCVD (first occurrence of nonfatal MI or CHD death, or fatal or nonfatal stroke) | 24626 | NR | NR | 2689 | 40-79 | 43.6% | NR | NR | Age, Total Cholesterol, HDL, SBP, Diabetes, Smoking |
| 25169175 | Randomly Selected Copenhagen residents without prior CVD events | 10-Year fatal CVD | 6991 | 1976 - 1978 | 11.9 | 2236 | 69.7 | 41.2% | 5.1% | 46.5% | Age, SBP, Total Cholesterol, Sex, Smoking, Diabetes, ECG Changes (Q waves, ST-segment depression, T-wave changes, ventricular conduction defects, LVH) |
| 25169175 | Randomly Selected Copenhagen residents without prior CVD events | 10-Year fatal or nonfatal CVD | 6991 | 1976 - 1978 | 9.8 | 3849 | 69.7 | 41.2% | 5.1% | 46.5% | Age, SBP, Total Cholesterol, Sex, Smoking, Diabetes, ECG Changes (Q waves, ST-segment depression, T-wave changes, ventricular conduction defects, LVH) |
| 1985385 | Members of the Framingham Heart Study and Framingham Offspring Study cohorts, without baseline CVD or cancer (other than basal cell carcinomas). | 10-Year CHD (MI and CHD death plus angina pectoris and coronary insufficiency) | 5573 | 1968 - 1975 | 12 | NR | 30-74 | 46.5% | 6.0% | 39.7% | Gender, age, SBP, smoking, total cholesterol, HDL, diabetes, LVH on EKG |
| 1985385 | Members of the Framingham Heart Study and Framingham Offspring Study cohorts, without baseline CVD or cancer (other than basal cell carcinomas). | 10-Year MI (including silent and unrecognized) | 5573 | 1968 - 1975 | 12 | NR | 30-75 | 46.5% | 6.0% | 39.7% | Gender, age, SBP, smoking, total cholesterol, HDL, diabetes, LVH on EKG |
| 1985385 | Members of the Framingham Heart Study and Framingham Offspring Study cohorts, without baseline CVD or cancer (other than basal cell carcinomas). | 10-Year CHD Mortality (including sudden and nonsudden) | 5573 | 1968 - 1975 | 12 | NR | 30-74 | 46.5% | 6.0% | 39.7% | Gender, age, SBP, smoking, total cholesterol, HDL, diabetes, LVH on EKG |
| 1985385 | Members of the Framingham Heart Study and Framingham Offspring Study cohorts, without baseline CVD or cancer (other than basal cell carcinomas). | 10-Year Stroke (including transient ischemic attack) | 5573 | 1968 - 1975 | 12 | NR | 30-74 | 46.5% | 6.0% | 39.7% | Gender, age, SBP, smoking, total cholesterol, HDL, diabetes, LVH on EKG |
| 1985385 | Members of the Framingham Heart Study and Framingham Offspring Study cohorts, without baseline CVD or cancer (other than basal cell carcinomas). | 10-Year CVD (MI, CHD death plus angina pectoris and coronary insufficiency, congestive heart failure, and peripheral vascular disease) | 5573 | 1968 - 1975 | 12 | NR | 30-74 | 46.5% | 6.0% | 39.7% | Gender, age, SBP, smoking, total cholesterol, HDL, diabetes, LVH on EKG |
| 1985385 | Members of the Framingham Heart Study and Framingham Offspring Study cohorts, without baseline CVD or cancer (other than basal cell carcinomas). | 10-Year CHD Mortality (including sudden and nonsudden) | 5573 | 1968 - 1975 | 12 | NR | 30-74 | 46.5% | 6.0% | 39.7% | Gender, age, SBP, smoking, total cholesterol, HDL, diabetes, LVH on EKG |
| 1985385 | Members of the Framingham Heart Study and Framingham Offspring Study cohorts, without baseline CVD or cancer (other than basal cell carcinomas). | 10-Year CHD (MI and CHD death plus angina pectoris and coronary insufficiency) | 5573 | 1968 - 1975 | 12 | NR | 30-74 | 46.5% | 6.0% | 39.7% | Gender, age, DBP, smoking, total cholesterol, HDL, diabetes, LVH on EKG |
| 1985385 | Members of the Framingham Heart Study and Framingham Offspring Study cohorts, without baseline CVD or cancer (other than basal cell carcinomas). | 10-Year MI (including silent and unrecognized) | 5573 | 1968 - 1975 | 12 | NR | 30-74 | 46.5% | 6.0% | 39.7% | Gender, age, DBP, smoking, total cholesterol, HDL, diabetes, LVH on EKG |
